# Digital markers from smartwatch data relate to non-motor clinical examinations of Parkinson’s disease

**DOI:** 10.1101/2023.09.12.23295406

**Authors:** Ann-Kathrin Schalkamp, Neil A Harrison, Kathryn J Peall, Cynthia Sandor

## Abstract

Monitoring of Parkinson’s disease (PD) has seen substantial improvement over recent years as digital sensors enable a passive and continuous collection of information in the home environment. However, the primary focus of this work has been motor symptoms, with little focus on the non-motor aspects of the disease. To address this, we combined longitudinal clinical non-motor assessment data and digital multi-sensor data from the Verily Study Watch for 85 participants from the Parkinson’s Progression Monitoring Initiative (PPMI) cohort with a diagnosis of PD. We show that digitally collected physical activity and sleep markers do significantly relate to clinical non-motor assessments of cognitive, autonomic, and daily living impairment. The observed poor predictive performance, however, highlights the need for better targeted digital markers to enable a monitoring of non-motor symptoms.

## Introduction

Though classified as a motor disorder, Parkinson’s disease (PD) is associated with multiple non-motor symptoms that frequently arise prior to clinical diagnosis and progress throughout the disease course (Chaudhuri et al., 2006; Schapira et al., 2017). Non-motor symptoms include psychiatric symptoms, autonomic and sleep disturbance, pain, fatigue, and cognitive impairment, with many recognised to impact quality of life to a greater extent than the motor symptoms associated with PD (Chaudhuri et al., 2006; Martinez-Martin et al., 2011).

Advances in digital heath technologies (DHTs) have enabled the development of sensors that can continuously and passively monitor symptoms outside of a clinical setting. Several DHTs have already been developed to track motor signs and symptoms in PD with the digital scores developed providing a good representation of the existing gold standard clinical rating scale, the Unified Parkinson’s Disease Rating Scale (UPDRS) (Bot et al., 2016; Burq et al., 2022; Lipsmeier et al., 2022; Powers et al., 2021; Sieberts et al., 2021). By contrast, apart from sleep, non-motor symptoms have been largely neglected in the context of DHTs (Li et al., 2023). Digital markers for sleep length, quality, and stage exist (Djanian et al., 2022), however, are rarely used for the monitoring of PD. In the few studies that do study digital and non-motor markers for PD, a link between digitally tracked bradykinesia and self-reported disturbed sleep (McGregor et al., 2018), and between digitally tracked bradykinesia and constipation (van Wamelen et al., 2019) were identified.

Here, we used rich multi-modal data from the Parkinson’s disease Progression Marker Initiative (PPMI) cohort to investigate how standard markers of physical activity, sleep, and vital signs obtained from passively collected free-living smartwatch data relate to clinically assessed non-motor signs and symptoms and evaluated their potential utility in the context of clinical care.

## Results

### Digital weekly averages in the PPMI cohort

At the time of data retrieval (November 2022), the PPMI dataset provided a mean of 485 days of at home monitoring for 14 features describing physical activity (step count, walking minutes), sleep (total time, REM time, NREM time, deep NREM time, light NREM time, wake after sleep onset (WASO), awakenings, sleep efficiency), and vital signs (pulse rate, mean root mean squared successive differences (RMSSD) (heart beat), median RMSSD, RMSSD variance) for 149 participants with a diagnosis of PD. Clinical data including assessments of cognitive performance, autonomic functioning, psychiatric symptoms, impairment in daily living, and motor symptoms was collected between 2010 and 2021 (Marek et al., 2018). A temporal overlap between the timing of the clinic visits and collection of the digital data was found for 85 subjects with a mean of 1.58±0.78 clinic visits per participant during the digital data collection period. PD cases were recruited no more than 2 years after their initial PD diagnosis such that at the last visit to the clinic coinciding with the digital data collection 6.81±2.11 years had passed since diagnosis, leading to a cohort of individuals diagnosed with PD with an average age of 67.69±7.54.

### Digital markers capture variability in motor and non-motor clinical assessments

Multiple groups of non-motor signs and symptoms demonstrated relatedness within their individual domains including cognitive (17 of 28 pairs reached significance after FDR correction), psychiatric (3 of 6), autonomic (1 of 6), and daily functioning (1 of 1) (Figure 2, Supplemental Table 1). Several of the non-motor signs and symptoms related to higher motor impairment (UPDRS II): reduced cognitive performance (2 of 8), increased RBDSQ (p-value = 9.6×10^-3^), reduced independence in daily functioning (2 of 2), and higher UPDRS IV (p-value = 1.34×10^-4^). The non-motor assessments were further related to independence in daily living and UPDRS IV. Psychiatric assessment demonstrated correlation with increased difficulties in daily living (6 of 8) and with increased UPDRS IV (GDS: p-value = 4.72×10^-2^, STAI trait: p-value = 1.93×10^-2^). Higher levels of autonomic dysfunction were shown to be associated with increased difficulty in daily living and SCOPA autonome (p-value = 8.01x 10^-3^) and RBDSQ (p-value = 1.54x 10^-4^) were additionally related to UPDRS IV. Few relationships to cognitive performance were identified with only higher MoCA scores correlated with smaller drop in systolic blood pressure (p-value = 3.78x 10^-2^).

There was also evidence of interrelatedness across digital markers including sleep (10 of 28), vital signs (1 of 3), and physical activity (1 of 1), although there was no evidence of a relationship between the domains.

The weekly averages of the digital markers (a 3.5-day window spanning the clinical visit date, Figure 1) correlated with several of the non-motor clinical markers for those diagnosed with PD (Figure 2, Supplemental Table 1). Of the eight cognitive markers, two were represented by the digital marker for step count (LNS: p-value = 6.35x 10^-4^, and symbol digit: p-value = 8.64×10^-3^) and four related to digital sleep measures; MoCA (p-value = 7.64×10^-3^) and HVTL recall (p-value = 2.07×10^-3^) were represented by total sleep time, LNS was negatively related to WASO (p-value = 1.93×10^-2^) and HVLT retention was positively related to light NREM sleep time (p-value = 4.37×10^-2^). Of the four autonomic functioning markers, two were captured by digital markers; SCOPA autonome was represented by WASO (p-value = 1.14×10^-2^) and ESS by both physical activity markers (p-value < 4.64×10^-2^). One of the two daily living questionnaires was captured through the digitally measured step count (Schwab England ADL: p-value = 8.01×10^-3^). High UPDRS IV scores were associated with increased WASO (p-value = 2.09×10^-2^). None of the psychiatric markers were captured by any of the digital timeseries components. A link between the digital and clinical motor markers was also observed with UPDRS III OFF being negatively related to step count (p-value = 3.54×10^-2^).

**Figure 1:**
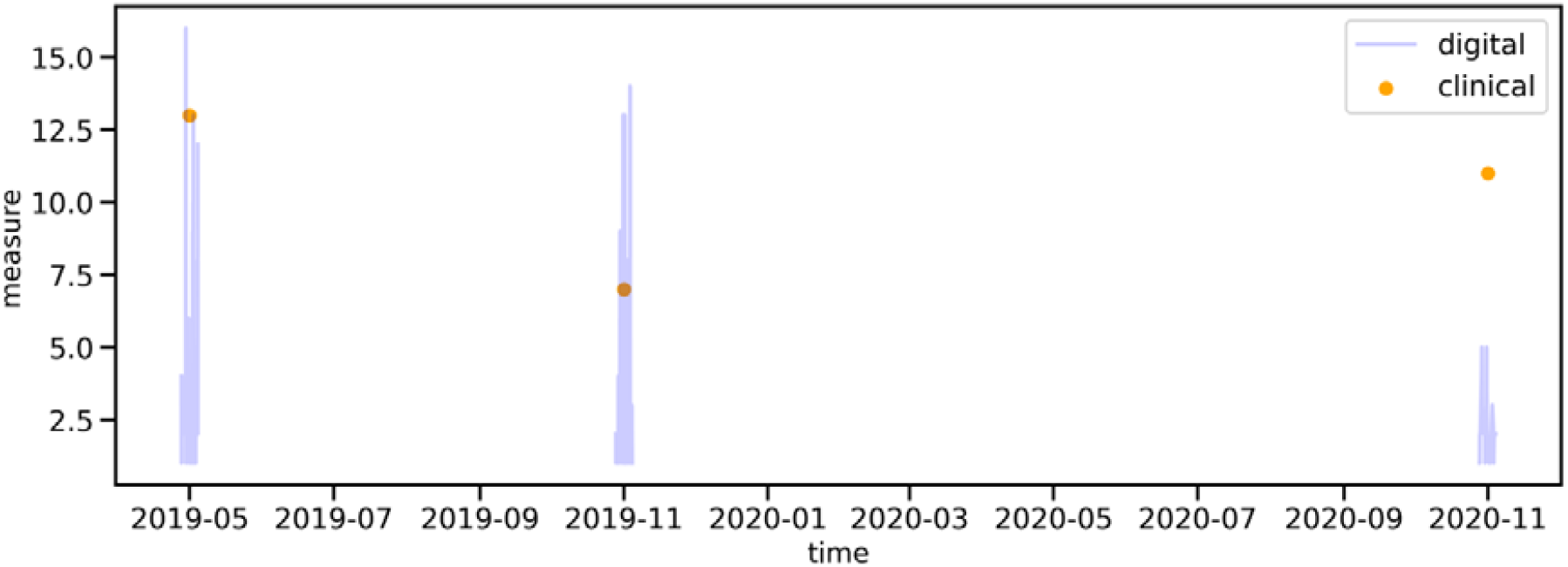
Deriving digital weekly averages. The continuously collected digital data was aligned with the visits to the clinic. An average of the digital markers around the clinic visit were calculated using a 3.5-day window excluding the day of the clinic visit itself, such that an average over 6 days was computed.

**Figure 2:**
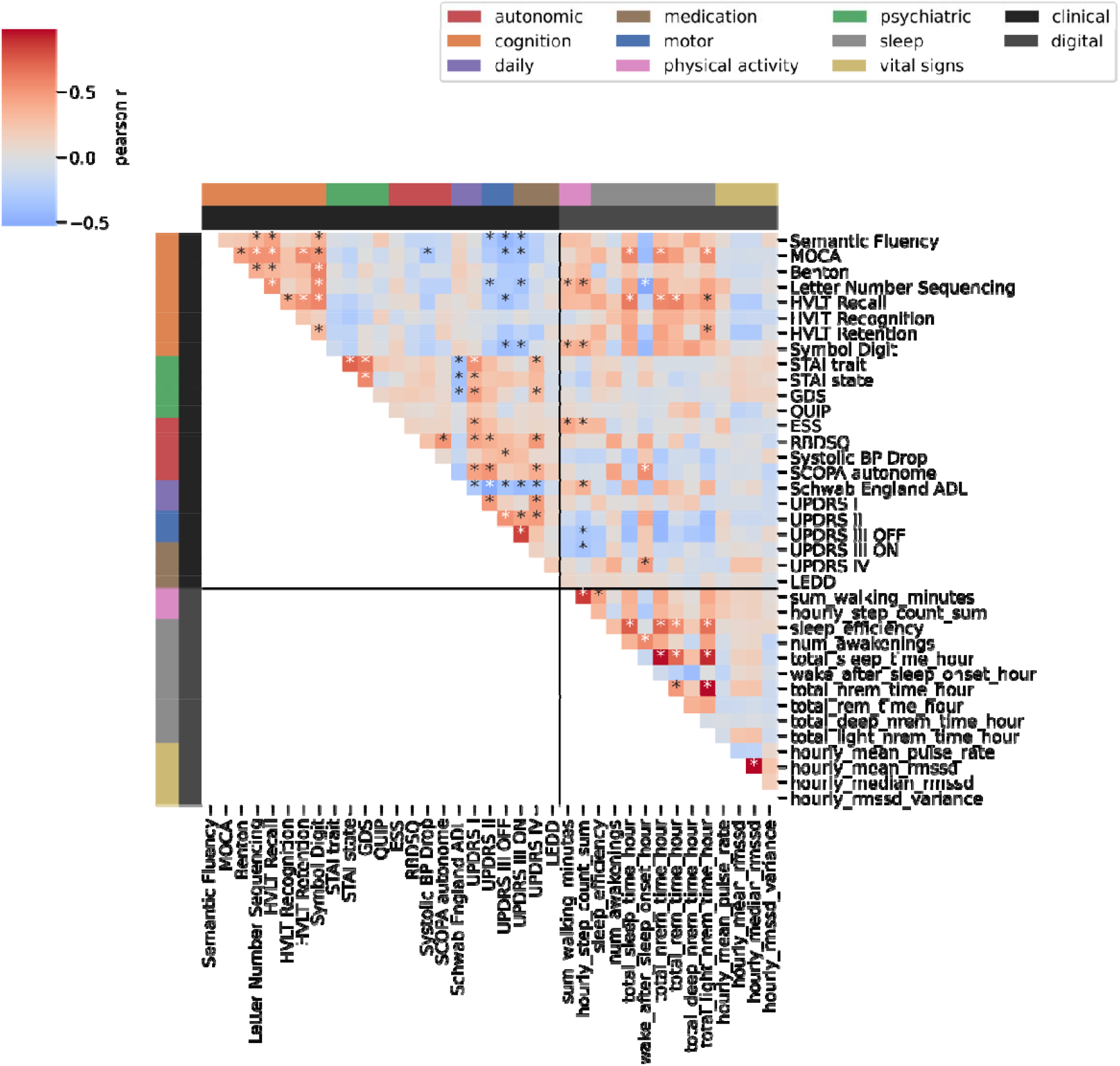
Correlation of digital weekly averages and clinical assessments. The heatmap displays the Pearson’s r coefficient for each digital marker and clinical assessment in the Parkinson’s disease group. If multiple visits overlapped with the digital data per person, the last visit to the clinic was chosen. Individual tests are grouped into modalities as indicated by the colours on the left and top. Asterisks indicate significant correlation after 0.05 FDR correction.

### Digital averages could not predict non-motor assessments on an individual level

While there were associations between digital and non-motor clinical markers, weekly digital averages could not predict the outcomes of standardised non-motor symptom questionnaires or rating scales used during the clinical assessments, on an individual level (Figure 3, Supplemental Table 2). Most models achieved an R2 below 0, indicating that no variation in the clinical data could be explained through the digital markers. Of note, the clinical motor markers were also not explained through the digital weekly averages. UPDRS II (R2 = 0.05±0.02, p-value = 1.83×10^-3^) and ESS (R2 = 0±0.04, p-value = 1.9×10^-3^) were the only markers that showed a significant improvement in performance compared to baseline models, which were trained on age, education, and male sex.

**Figure 3:**
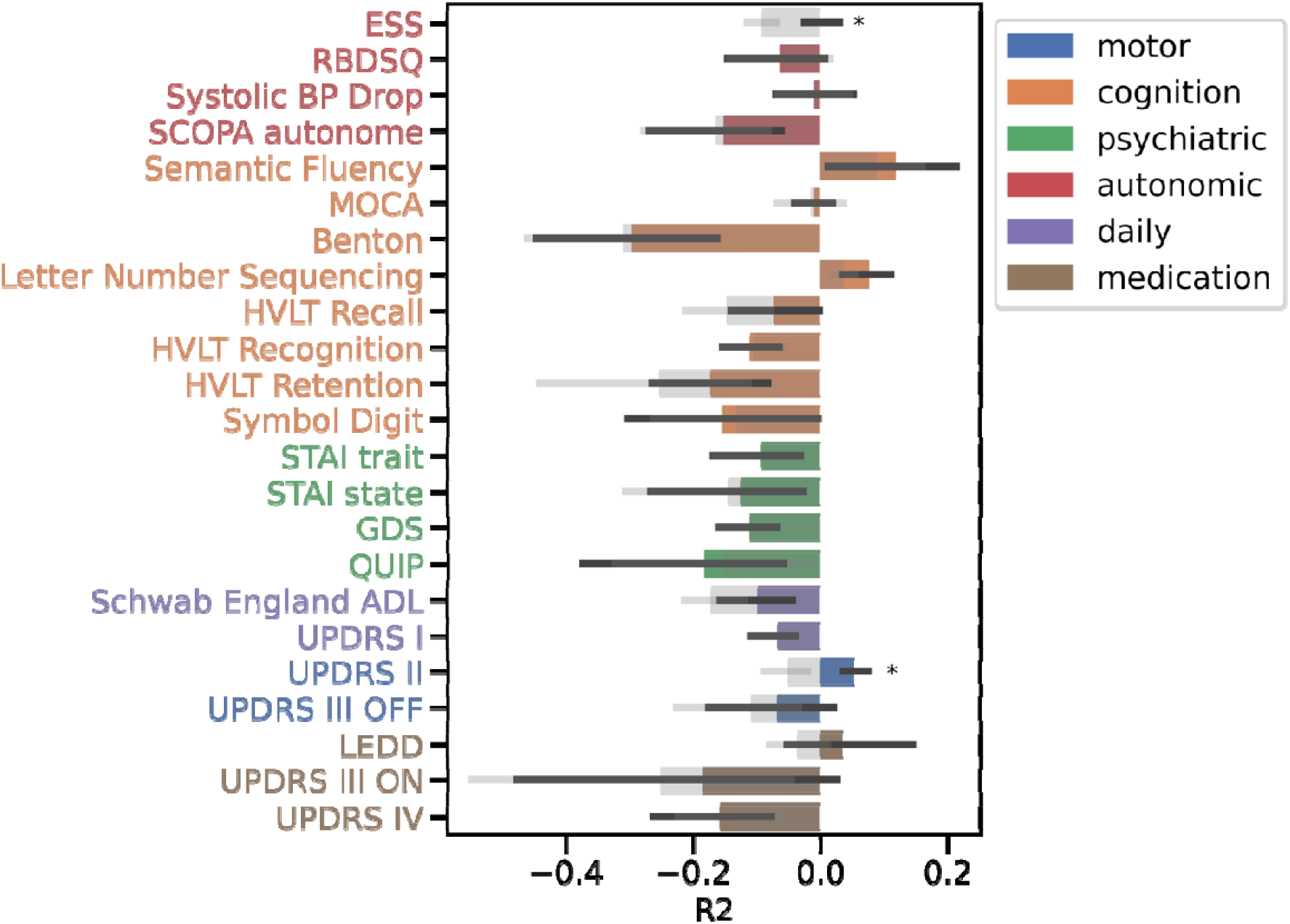
Digital markers fail to predict clinical markers. The mean R2 across the five outer cross-validation folds with their 95% Confidence Interval are shown. The barplots are overlayed with the baseline performance in grey also with 95% CI. An asterisk indicates significant improvement over baseline at 0.05 significance. The colour indicates the domain of the clinical assessment.

### Change in clinical non-motor features over time is not related to change in digital features over time

We computed the difference of the clinical rating scales and questionnaire scores between clinical visits, as well as the change of the weekly digital averages between clinical visits (Figure 4, Supplemental Table 3), identifying no significant associations between the rate of change of the clinical and digital markers. The rate of change of clinical markers showed no association amongst themselves, by contrast the rate of change of the digital markers were observed to be related within domains, however, this was anticipated as they derive from one another.

**Figure 4:**
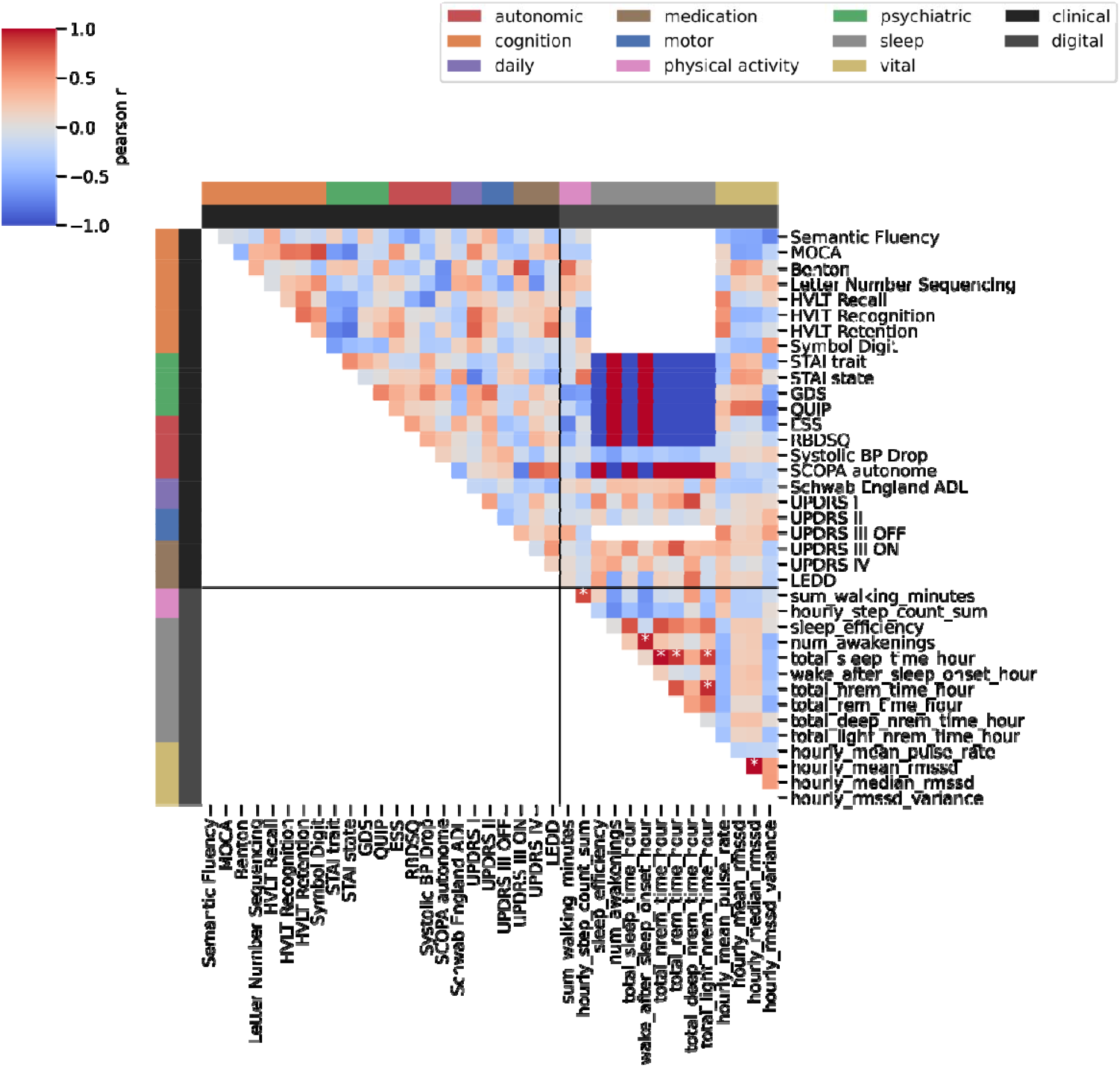
Rate of change in digital not related to rate of change in clinical. The heatmap displays for every combination of digital and clinical markers the correlation between the mean rate of change over time. The domains can be seen on the left and top coded by the colour bars. The colour indicates the Pearson r coefficient and an asterisk indicates 0.05 FDR corrected significance.

## Discussion

Here, we provide the first demonstration of how digital markers collected in free-living conditions may relate to non-motor clinical assessments in a PD cohort. More specifically, we show that measures capturing weekly averages of physical activity are concordant with clinical assessments of multiple non-motor symptoms including cognitive, autonomic, and daily functioning as well as motor signs and symptoms. However, digital markers demonstrated limited power in the prediction of the clinical scores, and the rate of change in digital markers did not relate to the rate of change in the clinical features.

While PD is primarily viewed as a motor disorder with symptomatic treatment focussing on relieve of such motor aspects, non-motor aspects of the disease can precede the onset of motor symptoms by several years and throughout the disease course can dominate the clinical presentation and impact reductions in quality of life (Schapira et al., 2017). While DHTs have proven successful in the passive and continuous monitoring of the motor symptoms of PD, few studies have sought to examine whether digital markers may also aid in monitoring and longitudinal understanding of the non-motor symptoms despite the recognition that this latter symptom group has the greater impact on quality of life in PD (Andrejack & Mathur, 2020; Martinez-Martin et al., 2011).

Previous studies have indicated the potential of DHTs in monitoring non-motor symptoms in PD with van Wamelen et al. (2019) reporting an association between digital bradykinesia scores and constipation. We found relations between step count and cognition in PD that were previously reported in other disorder like Schizophrenia (Chen et al., 2022) or subacute stroke (Ito et al., 2021). Our results further replicated the findings of digitally tracked poor sleep being related to worse cognitive performance in PD (Stavitsky et al., 2012) and self-reported sleep disturbances being associated with more PD treatment/motor complications (Antczak et al., 2013; Gomez-Esteban et al., 2006), which we identified with digitally measured increased WASO being related to increased UPDRS IV.

Our digital weekly averages were unable to predict clinical motor and non-motor markers on an individual level. Although previous research has shown that motor symptoms of PD (as captured with UPDRS) can be predicted from digital markers (Burq et al., 2022; Powers et al., 2021), digital sensor data collected in free-living conditions showed only few associations with the clinical motor severity scales (Galperin et al., 2019; Toosizadeh et al., 2015), which might be why we were unable to predict these motor symptoms from the digital weekly averages of standard markers obtained from smartwatches. It is likely that the here considered standard digital markers lack the specificity needed to accurately represent the clinical markers as they only describe high-level features of physical activity, sleep behaviour, and vital signs. More specific digital markers collected passively in real-life settings, such as those identified for tremor or dyskinesia (Powers et al., 2021), are currently lacking for the non-motor aspects of PD.

Limitations of this study relate primarily to data availability. Due to the Verily Study Watch only being introduced 10 years after the PPMI study began, the overlap between the clinical assessments and the digital data is limited to 85 participants who at the time of digital monitoring already had a diagnosis of PD for about seven years and received dopaminergic therapy. The sample size was even smaller for the rate of change analysis where at least two visits per person had to overlap with the digital data collection leading to only 35 subjects being considered in this analysis. Finally, our analysis was limited by only considering standard digital marker measures as provided by Verily, the code for which is proprietary, and therefore limiting the reproducibility of this work.

In conclusion, we have demonstrated associations between standard smartwatch digital markers and multiple non-motor aspects of PD including i) digitally measured sleep and cognition and motor complications due to medication, and ii) digitally measured physical activity and cognition, daytime sleepiness, and independence in daily living. Despite these associations, the clinical markers could not be predicted from digital data on an individual level, highlighting the need for more specialised digital markers for the non-motor aspects of PD.

## Supporting information

All Supplemental Tables

## Data Availability

All data used in this study was accessed from https://www.ppmi-info.org/accessdata-specimens/download-data. The Institutional Review Board approved the PPMI program and all participants gave written informed consent. For up-to-date information on the study, visit www.ppmi-info.org.

## Code Availability

All associated code to reproduce the analyses performed here will be made publicly available upon publication.

## Acknowledgements

We thank all participants of the PPMI study, all investigators, and the Michael J. Fox Foundation.

## Author Contribution

A-K.S. and C.S. participated in designing the study, topic definition, and review of relevant studies. Machine learning models and statistical analyses were designed and implemented by A-K.S.. Figures and tables were done by A-K.S. with the support of C.S.. A-K.S. wrote the first draft. A-K.S., C.S., N.A.H., and K.J.P. contributed to subsequent versions of the manuscript. All authors critically reviewed the paper, all authors have a clear understanding of the content, results, and conclusions of the study and agree to submit this manuscript for publication. The corresponding author (C.S.) declares that all authors listed meet the authorship criteria and that no other authors involved in this study are omitted. C.S. is ultimately responsible for this article.

## Methods

All analyses were performed in python v3.9 using sklearn (Pedregosa et al., 2011) 1.2.1 for model training and evaluation, scipy 1.10.0 and pingouin (Vallat, 2018) 0.5.3 for statistical testing, and matplotlib 3.6.3 and seaborn 0.12.2 for creating figures. Data loading and manipulation has been facilitated through an adapted version of pypmi (https://github.com/rmarkello/pypmi). All code will be made available upon publication.

### Study Cohort

The Parkinson’s disease progression marker initiative (PPMI) has collected data from those with recently diagnosed (*denovo*) PD, people at risk, and unaffected controls since 2010. They put a focus on longitudinal data collection of brain imaging, blood, urine, cerebrospinal fluid (CSF), and clinical assessment data. Since 2018 a subset of participants has been supplied with a Verily Study Watch, which is equipped with a multitude of sensors including accelerometer, gyroscope, electroencephalography (ECG), and photoplethysmography (PPG). We used the analytic dataset cohort assignment, which provides the most up-to-date assignment of subjects into PD, healthy control, Scans Without evidence of dopaminergic deficit (SWEDD), and prodromal class.

#### Digital Data

From the raw Verily Study Watch data several derived markers are provided by Verily and were accessed by us through the LONI website of PPMI in November 2022. These include data on sleep, physical activity, and vital signs. Each derived digital marker has a different availability with the sleep markers being the scarcest due to them only being available for hours spent asleep. The hourly step count data covers an average of 1.25 years (std = 0.54) with a mean recorded time of 0.91 years (std = 0.52). The time between the measurements ranges from one hour to 6.76 days with a mean of 137.56 minutes (std = 530.76). The sleep data covers an average of 1.19 years (std = 0.57) with a mean covered time of 8.4 days (std = 6.68). Here, the time between measurements is larger on average with 6.35 days (std = 19.83).

### Clinical data

Data was downloaded from LONI PPMI in 2021. The following clinical assessments were retrieved: motor assessments including UPDRS scores (part II, III), cognitive assessments like Montreal Cognitive Assessment (MoCA), semantic fluency, benton judgement of line orientation, and WMS-III Letter-Number Sequencing Test (LNS), Hopkins Verbal Learning Test (HVLT), and symbol digit, psychiatric questionnaires including State-trait-anxiety inventory (STAI), geriatric depression score (GDS), and Impulsive-Compulsive Disorders in PD (QUIP), as well as autonomic assessments like Scale for Outcomes in Parkinson’s disease for Autonomic Symptoms (SCOPA), Epworth Sleepiness Score (ESS), REM sleep behavior disorder screening questionnaire (RBDSQ), and blood pressure drop, and assessments of daily functioning with the modified Schwab England Activities of Daily Living (Schwab England ADL) and the UPDRS I. Information on medication was included as Levodopa equivalent daily dosage (LEDD) and UPDRS IV.

These data were collected, cleaned, and merged based on the subject identifier and visit date. For UPDRS III we distinguished between ON and OFF assessments with OFF being those where the subject was not on medication either due to not taking medication or because the medication was deliberately not taken for this assessment, and ON being all assessments conducted when the subject took the normal medication.

### Temporal Alignment

The local date for the derived digital markers was calculated based on the provided age in seconds, the weekday, and the local time. The age in seconds was transformed to a date using the date of birth. The weekday of this estimated date was compared to the provided weekday. If they did not match, the estimated date was shifted to the closest date which has the correct weekday. Using this estimated date, the digital data was merged to the clinical data. Due to the smartwatch study only being included later in the study, the overlap with the clinical examinations is limited. 85 participants with PD had an overlap of digital data and a clinic visit with a mean of 1.58±0.78 clinical visits per person during digital data collection.

### Correlation of clinical markers and digital weekly averages

From the digital timeseries data, the weekly average around the clinical visit was calculated (Figure 1). For this, the clinical visit date was used as the mid-week point around which a 3.5-days sized window of the digital data was averaged, removing the visit day itself due to it being a non-representative day including a visit to the clinic. Thus, a mean over 6 days of all available data is calculated. We restricted this analysis to only include each participant to avoid overrepresentation of specific subjects by choosing the last available clinical visit with an overlapping digital recording available. The computed digital averages were correlated with the clinical visit information with FDR correction. Due to varying availability of clinical assessments, the number of subjects in each correlation differs.

### Prediction of clinical markers from digital weekly averages

To predict the clinical markers from the digital, we built regression models using all digital weekly averages as predictors and diagnosis age, time since diagnosis, and male as covariates. To estimate the baseline performance, a model using only the covariates was built. All these models used an elastic net penalty with R2 score loss and were fitted in a nested five-fold cross-validation. In the inner split gridsearch was applied to identify the best hyperparameters for the penalty, namely the L1 to L2 ratio and the alpha (strength of penalty) parameter. Performance was reported with mean and standard deviation of the R2 score across the five outer folds. Feature importance was investigated as the assigned coefficients across the outer folds being significantly different from zero (three standard deviations).

### Estimate rate of change

To assess whether the rate of change of the digital markers related to the change in the clinical markers, we computed the difference between visits scaled by the time between visits. 35 subjects had more than 2 overlaps of the digital data and a clinic visit. The mean over this change over time was correlated between clinical and digital data with FDR correction.

