## Supplemental Legends for "Digital markers from smartwatch data relate to non-motor clinical examinations of Parkinson’s disease"

Supplemental Material

Supplemental Table 1:
Results of the Pearson correlation analysis between clinical and digital markers. The Pearson's r coefficient, the associated p-value, the FDR corrected p-value, and the sample size is displayed.

Supplemental Table 2:
Results of the predictive modelling of clinical scores from digital weekly averages. For each clinical score the baseline R2 performance and the performance of the model using the digital markers as features are shown as mean R2 and standard deviation. The results of the two-sided T-test comparing the 5-fold cross-validation R2 performances between baseline and fitted model are shown with the t statistic and the associated p-value.

Supplemental Table 3:
Results of the Pearson correlation analysis between the rate of change in clinical and in digital markers. The Pearson's r coefficient, the associated p-value, the FDR corrected p-value, and the sample size is displayed.
